# Association of ACE I/D Polymorphism with Essential Hypertension in South-Asian Populations: Gender-Bias in Indian Populations—Evidence from Case-Control and Meta-Analysis Studies

**DOI:** 10.1101/2025.08.22.25334212

**Authors:** Shreya Sopori, Sonali Bhan, Arti Dhar, Audesh Bhat

**Author notes:** Correspondence: Dr. Audesh Bhat, Central University of Jammu-181143.

## Abstract

**Introduction:** The role of Angiotensin Converting Enzyme *(ACE)* intron-16 I/D polymorphism (rs4646994) in essential hypertension (EH) is contradictory in South Asian populations. The study objectives were to test the correlation of rs4646994 polymorphism with EH in the North-Indian population of Jammu and to perform a meta-analysis to validate its role in South-Asian populations.

**Methodology:** A total of 422 cases and 395 controls were genotyped in the original analysis. Pooled analysis was performed on 4987 cases and 5302 controls following the PRISMA and STREGA guidelines. SPSS v25.0 and web-based tools were used for statistical analyses.

**Results:** Our original case-control study revealed a statistically significant association between the rs4646994 polymorphism and EH, with the D allele being the risk allele. The observed significance level was higher when the recessive genetic model was tested [OR=1.68(95%CI:1.16–2.43), P=0.006]. Interestingly, only males carrying the DD genotype were at significantly higher odds of developing EH [OR=2.68(95%CI:1.58–4.53), P<0.0001]. The meta-analysis further corroborated an increased risk for EH in the presence of DD genotype in the South-Asian populations [OR=1.48 (95%CI:1.35–1.62), P<0.0001] and largely supported the gender-wise differences in Indian populations, with males at relatively higher risk.

**Conclusion:** Our results provide strong evidence supporting the role of rs4646994 polymorphism in EH in South-Asian populations, particularly Indians.

## Introduction

Essential hypertension (EH), also known as primary hypertension, is a predominant and highly prevalent type of hypertension, a medical condition of consistently elevated arterial blood pressure (BP) of ≥140 mmHg systolic and ≥90mmHg diastolic BP as per the WHO guidelines. In the last three decades, particularly in developing and underdeveloped countries, EH has become a major health problem, with the number of cases almost doubled, affecting both men and women almost equally [1]. Being a major risk factor for cardiovascular diseases (CVDs), kidney damage, and stroke, all leading causes of premature death, understanding the genetic and non-genetic causes is important for better management of this asymptomatic life-threatening condition [1]. The etiology of EH is often complex, involving interaction between multiple genetic and non-genetic factors [2], with data supporting a cumulative effect of multiple single nucleotide polymorphisms (SNPs) on BP increase rather than a single SNP effect [3, 4]. Smoking, stress, excessive alcohol intake, lack of physical activity, and certain dietary choices, such as high NaCl and low potassium intake, are some of the well-known non-genetic factors. In contrast, the genetic factors include around 900 BP-related nucleotide variants in different genes, including the Renin-Angiotensin-System (RAS) and circadian rhythm genes [5-13].

RAS is the main regulator of BP, sodium metabolism, vascular modeling, and renal hemodynamics, therefore, variation in BP may result from the genetic changes in any of RAS controlling components [8, 14] Concordantly, several genetic variations in the RAS pathway genes; angiotensinogen (*AGT*), angiotensin I-converting enzyme (*ACE*), renin (*REN*), angiotensin-II type-1 receptor (*AT1-R*), and angiotensin-II type-2 receptor (*AT2-R*) have been found associated with HTN [15-17]. ACE, a zinc metallopeptidase, considered as the primary enzyme of RAS pathway, converts angiotensin-I to angiotensin-II which in turn attaches to plasma membrane receptors, producing arteriolar constriction and a surge in diastolic and systolic BP [2]. ACE is encoded by a 21 kb *ACE* gene located on chromosome 17 (17q23-q24) having 26 exons and 25 introns [18-23]. An insertion (I)/deletion (D) polymorphism of 287-bp Alu repeat in the intron-16 of this gene (rs4646994/rs1799752) has been associated with variable serum ACE levels, with the ‘D’ allele having the maximum expression levels [24-33]. Published literature suggests a conflicting role for this polymorphism in human diseases, including EH [2, 34-38]. Likewise, data from South Asian populations even though largely supporting a positive correlation between the D allele and EH, inconsistencies do exist in the literature, with many studies either showing a lack of correlation [2, 20, 24, 28, 29, 37, 39-41] or a negative correlation between the D allele and HTN [42] (**Suppl. Table 1**).

Considering the significant risk factor *ACE* I/D polymorphism has emerged for EH in some Indian populations, we first studied its potential role in EH in the North Indian population of Jammu region, a hitherto unstudied population. This was followed by a meta-analysis on the pooled data from South Asian populations and our original study to validate the role of this polymorphism in EH in the South Asian populations and, importantly to evaluate if gender-specific variance in susceptibility exists in the South Asian populations in general.

## Materials and Methods

### Case-control study

#### Ethical statement

The Institutional Human Ethical Committee (IHEC) of the Central University of Jammu approved this study vide notification no. IHEC/CUJ/CMB-23/01. Participants gave their informed consent to participate in this study. Relevant details from the participants were entered into a predesigned questionnaire. All experiments and data collection were carried out in compliance with the IHEC norms.

#### Subjects

In total, 817 subjects (422 cases and 395 ethnicity, age, and gender-matched healthy controls) were enrolled from the Jammu region of UT of Jammu and Kashmir. Ethnicity was confirmed before enrollment in the study. A statistical comparison of demographic characteristics between cases and controls is presented in **Table 1**. Subjects suffering from or having any family history of secondary HTN, diabetes, renal disorders, thyroid problem or CVDs were excluded from the study. Normotensive subjects with any family history of EH were also excluded from the control group. After 5 mins rest in a seated position post arrival at the sample collection centers (Government Medical College Jammu and Rama Krishna Medical Centre, Jammu), three independent BP measurements each at an interval of 5-10 mins were taken from the participants by a trained healthcare professional using standard Sphygmomanometer. The average difference in systolic blood pressure (SBP) of cases and controls was statistically significant (155.24±12.6 mmHg vs. 115.50±8.5 mmHg, P<0.0001). Likewise, the diastolic blood pressure (DBP) was also statistically different with the average DBP of cases 89.35±8.7 mmHg and controls 72.41±6.1 mmHg (P<0.0001). The difference in the proportions of females and males in cases and controls was found statistically non-significant (P=0.616). A post-hoc power analysis using QUANTO program yielded a >80% study power.

**Table 1:** Demographic characteristics and clinical parameter of the controls and cases.

| **Characteristics** | **Controls (N=395)** | **Cases (N=422)** | **P value** |
| --- | --- | --- | --- |
| Age (Yrs) | 54.15±12.6 | 55.90±12.5 | 0.05 |
| BMI | 24.56±3.7 | 24.45±4.4 | 0.692 |
| Gender | Female (n=157) | Female (n=175) | 0.616 |
|  | Male (n=238) | Male (n=247) |  |
| SBP | 115.50±8.5 | 155.24±12.6 | ≤0.0001 |
| DBP | 72.41±6.1 | 89.35±8.7 | ≤0.0001 |

BMI, Body Mass Index; DBP, Diastolic Blood Pressure, SBP, Systolic Blood Pressure; Yrs, Years. Unpaired t-test was performed to calculate the probability (p) values

#### DNA extraction and genotyping

The genomic DNA extraction protocol was previously described [13]. For genotyping the *ACE* I/D polymorphism, we used previously reported I/D flanking primer pair with some modification. After performing In-silico PCR (https://genome.ucsc.edu/cgi-bin/hgPcr) to confirm the target specificity, we detected deletion of G nucleotide in the reported forward primer (highlighted in bold and underlined) and a C>T transition in the reverse primer (highlighted in bold and underlined). The modified primers used in our PCR were; forward 5’-CTGGAGA**<u>G</u>**CCACTCCCATCCTTTCT-3’ and reverse 5’-GA**<u>C</u>**GTGGCCATCACATTCGTCAGAT-3’) [23]. A second primer pair having an insertion-specific forward primer 5’-TTTGAGACGGAGTCTCGCTC-3’ as reported by [23] and the above mentioned reverse primer was used to validate the insertion sequence to avoid mistyping of ID genotype as DD genotype. Polymerase chain reaction was performed in the presence of 5% DMSO as described previously. The same PCR conditions and reagents were used to amplify the target region of the *ACE* gene as reported previously [13], except that the annealing temperature of 1st primer pair was set at 63°C and 2nd pair at 61°C. The amplification products were subjected to electrophoresis on a 2% agarose gel to record the genotypes. Genotyping performed with primer pair 1 was recorded as follows: II = 490bp, DD = 190bp, ID = 490bp and 190bp bands. In an ID heterozygote, the I allele’s amplification is sometimes suppressed causing the latter to be mistyped as DD. Individuals who were typed as DD with the 1st primer pair were regenotyped with the 2nd primer pair to detect the presence of 408bp amplicon. II genotype sample was used as a positive control. Except for the annealing temperature, all PCR conditions were kept the same.

## Meta-analysis

### Literature search

To evaluate the relationship between *ACE* I/D polymorphism and susceptibility towards EH in South Indian populations, including the gender-wise differences, relevant published data was retrieved from PubMed, Google Scholar, ResearchGate, Web of Science, Scopus, Semantic Scholar, and The Lens databases. Literature search was performed by using the Boolean operators “AND” and “OR” to combine/connect at least one word from each block. Eligible studies were shortlisted using the Eligible Studies Criteria for Systematic Meta-Analyses (PRISMA) guidelines [43]. The following keywords were used for the literature search: Hypertension and *ACE* indel polymorphism; essential hypertension angiotensin converting enzyme indel polymorphism. These terms were searched in combination with South Asia, Bangladesh, Bhutan, India, Maldives, Pakistan, Sri Lanka, and Nepal. Using subject keywords plus free words in our search minimized the omission of relevant studies. The available language was restricted to English only. Articles were shortlisted first based on the title and abstract, followed by full-text assessment using our inclusion/exclusion criteria. Two independent reviewers assessed the quality of included studies using the PRISMA model (**Figure 1**).

**Figure 1:**
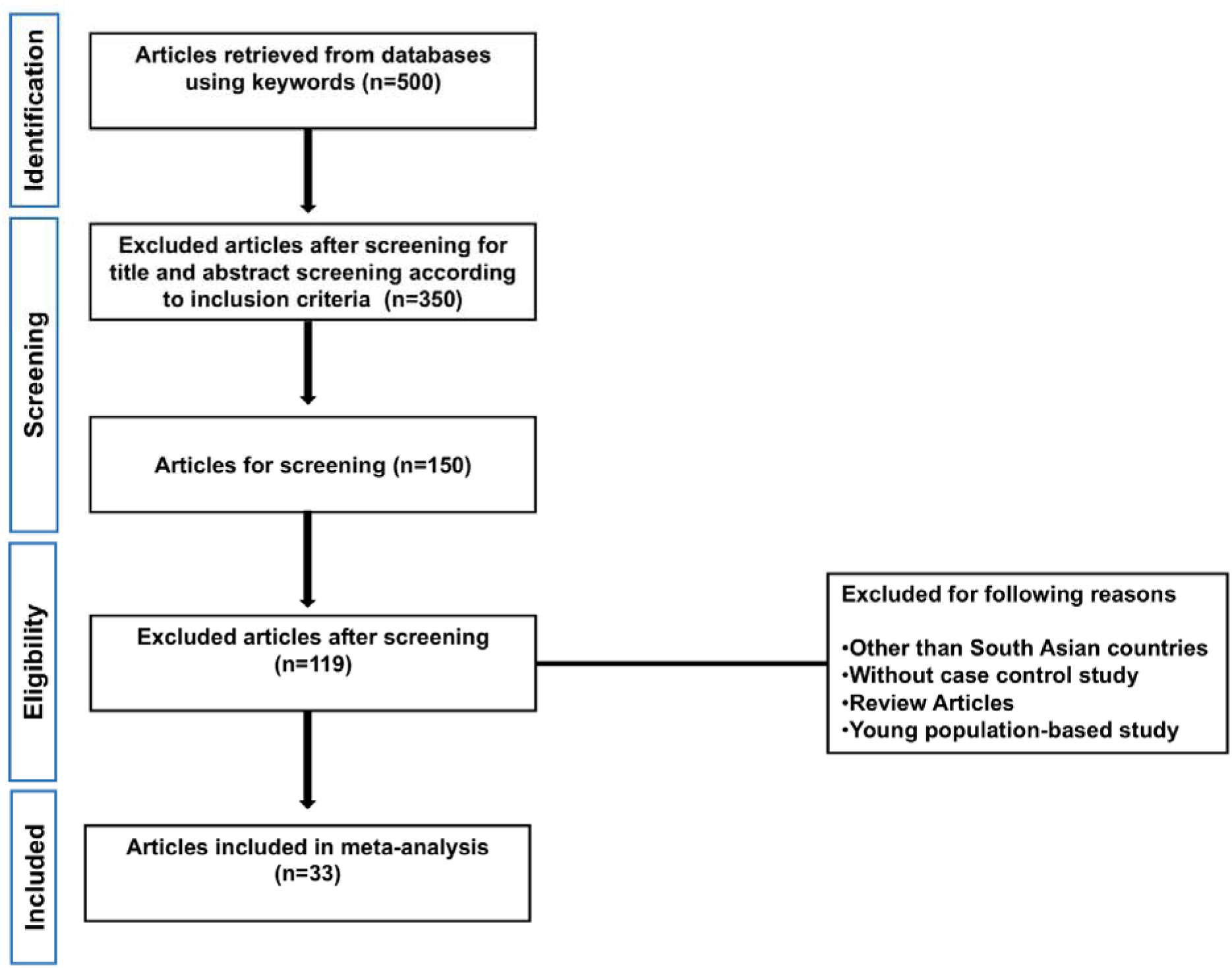
Flow diagram for selection of studies.

#### Inclusion/exclusion criteria and Data extraction

Studies were included if they were a case-control study; controls were healthy individuals without any comorbidity; cases represented only EH patients without any comorbidity; conducted on adult population (>19 years of age) of South Asian origin; availability of full-text. Reasons for exclusion included; if not a case-control study, was a systematic review or meta-analysis; not on the South Asian population; full-text not available, published in a non-indexed journal or a regional language. From each included study, three distinct kinds of data were extracted: basic information, primary study outcomes, and gender-specific data. The basic information included sample size (cases and controls), number of males and females, and average age, BMI, SBP, and DBP. Primary outcome data included distribution of genotype frequency and allelic frequency in cases and controls, and SBP and DBP subcategories if available. In gender-specific data, genotype/allelic frequency in males and females of cases and controls was extracted if available.

#### Quality check

We followed the 22-item Genetic Association Studies Report (STREGA) guideline [44] to evaluate the quality of the included studies. Each item was recorded as “yes” and “no” and was given a score of 1 for “yes” and 0 for “no”. Based on the total score earned, studies were ranked into high quality (15–22 score), medium quality (8–14 score), and poor quality (0–7 score) **(Suppl. File 1).** Two independent reviewers performed the quality check, with agreement in both considered as satisfactory. In case of disagreement, a third reviewer was involved. Hardy-Weinberg equilibrium test was performed on the available genotypic frequencies if not available in the original study.

### Statistical analysis

Numerical and categorical data such as age, gender, BMI, and BP of cases and controls were statistically compared with unpaired t-test using the web-based GraphPad tool (https://www.graphpad.com/quickcalcs/ttest1/). Differences in genotype frequencies between cases and controls and the Hardy□Weinberg equilibrium (HWE) in controls were assessed through Pearson’s chi-square (χ2) statistic (https://www.socscistatistics.com/tests/chisquare2/default2.aspx). The age, gender, and BMI adjusted odds ratio (OR) at 95% confidence interval was calculated via binary logistic regression using SPSS statistical software v25.0 (IBM, Armonk, NY, USA) to assess the level of association between the genotypes and EH. The II genotype (dominant) was coded as 2 (reference group), the ID genotype as 1, and the DD (recessive genotype) as 0. Three genetic models; codominant (II vs. ID, and II vs. DD), dominant (II vs. ID+DD), and recessive (DD vs. II+ID) were tested to measure the level of association between the I/D polymorphism and EH. Association statistics of our case-control study were further validated through the online statistical tool SNPStats (https://www.snpstats.net/start.htm). The statistical power of the variant was calculated via the Genetic Power Calculator (http://pngu.mgh.harvard.edu/~purcell/gpc/) [45] and was found >80%. For significance levels, a P value of <0.05 was considered significant.

## Results

### Case-control study

The allele and genotype frequencies of the *ACE* gene I/D polymorphism in cases and controls are given in **Table 2**. The polymorphism was found in HWE in the control group (P=0.26). Differences in the allele frequency distribution between cases and controls was found non-significant (P=0.66); however, the genotype distribution between cases and controls was found statistically different (P=0.005), with the DD genotype being more prevalent in cases than controls (21.8% vs. 13.9%) (**Table 2**). The logistic regression statistics revealed a significant association under the recessive genetic model (II+ID vs. DD) with an adjusted OR of 1.68 (95%CI: 1.16–2.43, adjusted P=0.006) and a slightly non-significant association under the co-dominant model (II vs. DD) with an adjusted OR of 1.47 (95%CI: 0.97–2.22, adjusted P=0.06). The statistical significance was found non-significant under the dominant model (II vs. ID+DD) (**Table 2**).

**Table 2:** Genotype and allele frequency distribution in cases and controls under different genetic models and their statistical significance.

| Genetic Model | Genotype | Genotype Frequency |  | Allelic frequency |  | HWE* | P value | OR<sup>#</sup>(95%CI) |
| --- | --- | --- | --- | --- | --- | --- | --- | --- |
|  |  | Controls (N=395) | Cases (N=422) | Controls | Cases |  |  |  |
| Codominant model | II | 135(34.2%) | 149(35.3%) | I = 0.6 | I = 0.57 | 0.26 | Reference | 1 |
|  | II vs ID | 205(51.9%) | 181(42.9%) | D = 0.4 | D = 0.43 |  | 0.14 | 0.79 (0.58-1.08) |
|  | II vs DD | 55(13.9%) | 92(21.8%) |  |  |  | **0.06** | **1.47 (0.97-2.22)** |
| Dominant model | II | 135(34.2%) | 149(35.3%) |  |  |  | Reference | 1 |
|  | II vs ID+DD | 260(65.8%) | 273(64.7%) |  |  |  | 0.67 | 0.94 (0.70-1.25) |
| Recessive model | II+ID | 340(86.1%) | 330(78.2%) |  |  |  | Reference | 1 |
|  | II+ID vs DD | 55(13.9%) | 92(22.8%) |  |  |  | **0.006** | **1.68 (1.16-2.43)** |

\*Hardy-Weinberg equilibrium (HWE) corresponds to controls. <sup>#</sup>Values adjusted for age, sex and BMI. Reference refers to the genotype(s) with which the other genotype(s) was/were compared to in order to calculate the OR (Odds ratio) and p values. Logistical Regression was used to calculate the OR and 95% CI (Confidence Interval) and Pearson's chi square test to calculate the HWE

To assess the subcategory-wise correlation between *ACE* I/D and EH, we performed subgroup analyses based on DBP, SBP, gender, and HTN stages (Stage-I and Stage-II). Since the number of subjects in the obese group (BMI ≥30) and younger age group (<45 years) were less, we excluded such subtype analysis from this study. We also excluded sub-analysis based on lifestyle factors, such as smoking and drinking, the two important non-genetic risk factors due to data reliability concerns as Indians normally tend to hide or provide incorrect data due to social reasons. The baseline characteristics of the included subcategories are given in **Suppl. Table 2**. Similar to the combined group outcomes, both SBP and DBP independently showed a statistically significant association under the recessive model with a calculated OR of 1.68 (95%CI: 1.16–2.42, adjusted P=0.005) and 1.52 (95%CI: 1.04–2.20, adjusted P=0.02), respectively (**Suppl. Table 3**). Under the codominant model (II vs DD) we observed a slightly non-significant association in the SBP category with an OR of 1.49 (95% CI 0.99–2.23, adjusted P=0.05), and a non-significant association in the DBP category (P=0.13) (**Suppl. Table 3**). Significance levels under the dominant model were found non-significant, similar to what was observed in the combined set (**Table 2 and Suppl. Table 3**). These observations suggest that the *ACE* I/D polymorphism is a risk factor for both SBP and DBP.

Our gender-wise association analyses revealed that males with DD genotype are at a ∼3-fold higher risk of developing EH [OR 2.53 (95%CI: 1.42–4.49), adjusted P=0.001] (**Table 3**). Likewise, DD genotype in males indicated a strong correlation with the elevated DBP and SBP (adjusted P=0.002 and <0.0001, respectively). In comparison, a similar analysis revealed no significant association in females, thus suggesting that the DD genotype is more likely a risk factor only in males of the studied population (**Table 3**).

**Table 3:** Gender-wise association with essential hypertension in the Jammu population.

| Genetic Model | Genotype | Males |  | P value | OR<sup>#</sup> (95%CI) | Females |  | P value | OR<sup>#</sup>(95%CI) |
| --- | --- | --- | --- | --- | --- | --- | --- | --- | --- |
|  |  | Controls | Cases |  |  | Controls | Cases |  |  |
| **Combined Data** |  | **N=238** | **N=247** |  |  | **N=157** | **N=175** |  |  |
| Codominant model | II | 88(36.9%) | 82(33.2%) | Reference | 1 | 47(29.9%) | 67(38.3%) | Reference | 1 |
|  | II vs ID | 127(53.4%) | 109(44.1%) | 0.62 | 0.90 (0.60-1.34) | 78(49.7%) | 72(41.1%) | 0.06 | 0.62 (0.37-1.02) |
|  | II vs DD | 23(9.7%) | 56(22.7%) | **0.001** | **2.53 (1.42-4.49)** | 32(20.4%) | 36(20.6%) | 0.36 | 0.75 (0.41-1.38) |
| Dominant model | II | 88(36.9%) | 82(33.2%) | Reference | 1 | 47(29.9%) | 67(38.3%) | Reference | 1 |
|  | II vs ID+DD | 150(63.1%) | 165(66.8%) | 0.45 | 1.15 (0.79-1.68) | 110(70.1%) | 108(61.7%) | 0.08 | 0.66 (0.41-1.05) |
| Recessive model | II+ID | 215(90.3%) | 191(77.3%) | Reference | 1 | 125(79.6%) | 139(79.4%) | Reference | 1 |
|  | II+ID vs DD | 23(9.7%) | 56(22.7%) | **<0.0001** | **2.68 (1.58-4.53)** | 32(20.4%) | 36(20.6%) | 0.96 | 0.98 (0.57-1.69) |
| **DBP** |  | **N=345** | **N=140** |  |  | **N=214** | **N=118** |  |  |
| Codominant model | II | 126(36.5%) | 44(31.4%) | Reference | 1 | 68(31.8%) | 46(38.9%) | Reference | 1 |
|  | II vs ID | 177(51.3%) | 59(42.2%) | 0.77 | 0.93 (0.59-1.47) | 100(46.7%) | 50(42.4%) | 0.23 | 0.73 (0.44-1.22) |
|  | II vs DD | 42(12.2%) | 37(26.4%) | **0.002** | **2.43 (1.38-4.27)** | 46(21.5%) | 22(18.6%) | 0.25 | 0.69 (0.36-1.30) |
| Dominant model | II | 126(36.5%) | 44(31.4%) | Reference | 1 | 68(31.8%) | 46(38.9%) | Reference | 1 |
|  | II vs ID+DD | 219(63.47%) | 96(68.6%) | 0.34 | 1.22 (0.80-1.86) | 146(68.2%) | 72(61.1%) | 0.17 | 0.72(0.45-1.15) |
| Recessive | II+ID | 303(87.8%) | 103(73.6%) | Reference | 1 | 168(78.5%) | 96(81.4%) | Reference | 1 |
| model | II+ID vs DD | 42(12.2%) | 37(26.4%) | **<0.0001** | **2.53 (1.54-4.16)** | 46(21.5%) | 22(18.6%) | 0.49 | 0.81(0.46-1.44) |
| **SBP** |  | **N=249** | **N=236** |  |  | **N=169** | **N=163** |  |  |
| Codominant | II | 93(37.3%) | 77(32.6%) | Reference | 1 | 51(30.1%) | 63(38.6%) | Reference | 1 |
| model | II vs ID | 131(52.6%) | 105(44.5%) | 0.79 | 0.94 (0.63-1.41) | 84(49.7%) | 66(40.5%) | 0.07 | 0.63 (0.39-1.04) |
|  | II vs DD | 25(10.1%) | 54(22.9%) | **0.001** | **2.52 (1.43-4.43)** | 34(20.1%) | 34(20.9%) | 0.44 | 0.79 (0.43-1.45) |
| Dominant | II | 93(37.3%) | 77(32.6%) | Reference | 1 | 51(30.1%) | 63(38.6%) | Reference | 1 |
| model | II vs ID+DD | 156(62.6%) | 159(67.4%) | 0.33 | 1.20 (0.82-1.75) | 118(69.9%) | 100(61.4%) | 0.10 | 0.68 (0.43-1.07) |
| Recessive | II+ID | 224(89.9%) | 182(77.11%) | Reference | 1 | 135(79.9%) | 129(79.1%) | Reference | 1 |
| model | II+ID vs DD | 25(10.1%) | 54(22.9%) | **<0.0001** | **2.59 (1.55-4.34)** | 34(20.1%) | 34(20.9%) | 0.94 | 1.02 (0.59-1.74) |

#Values adjusted for age, sex. Reference refers to the genotype(s) with which the other genotype(s) was/were compared in order to calculate the OR (Odds ratio) and p values. Logistical Regression was used to calculate the OR and 95% CI (Confidence Interval)

To understand any age-related correlation, we divided subjects into <45 and ≥45 years of age as described previously (12). In the overall sample set, ≥45 years group showed significant association under the recessive model with the calculated OR of 1.57 (95%CI: 1.05–2.35, P=0.02). The gender-wise age group analysis revealed DD males of ≥45 yrs age at statistically higher odds of developing EH, as revealed by the codominant (II vs. DD) [OR 2.38 (95%CI: 1.28–4.44), adjusted P=0.006] and recessive [OR 2.44 (95%CI: 1.39–4.29), adjusted P=0.002] models, whereas, in the females of this age group, a protective effect was observed under the codominant (II vs. ID) [OR 0.51 (95%CI: 0.29–0.91), adjusted P=0.02] and dominant [OR 0.56 (95% CI: 0.33–0.95), adjusted P=0.03] models **(Suppl. Table 4)**. The <45 age group on the other hand showed non-significant differences between cases and controls in both males and females (**Suppl. Table 4**). However, these non-significant differences could be due to a small sample size.

Our stage-wise analysis revealed a strong positive correlation with Stage-I EH in the combined sample set under the codominant (II vs. DD) [OR 1.66 (95%CI: 1.05–2.65), adjusted P=0.03] and recessive model [OR 1.73 (95%CI: 1.14–2.61), adjusted P=0.009]. On the contrary, Stage-II largely showed a non-significant association with ACE I/D (**Suppl. Table 5**). In the gender groups of Stage-wise analysis, a statistically significant association was observed in Stage-I males under the codominant (II vs. DD) and recessive model with an observed ORs of 2.93 (95% CI 1.55–5.52, P=0.001) and 2.81 (95%CI: 1.59–4.96, adjusted P<0.0001) respectively, whereas females showed non-significant differences (**Suppl. Table 5**). In Stage-II men, a significant association was observed only under the recessive model [OR 2.29 (95%CI: 1.16–4.50, adjusted P=0.01]. On the contrary, Stage-II females showed a negative correlation with *ACE* I/D in the codominant (II vs. ID) and dominant models [OR 0.44 (95%CI: 0.23–0.85), adjusted P=0.01] and [OR 0.50 (95%CI: 0.28–0.90), adjusted P=0.02, respectively] (**Suppl. Table 5**).

### Meta-analysis

Our case-control results convincingly suggested a role for this polymorphism in EH in the Jammu population, more importantly, the existence of gender-specific role. Given the mixed evidence regarding the association of this polymorphism with EH and the absence of gender-specific differences in South Asian populations, we conducted a meta-analysis to explore these possibilities in the existing literature.

#### Selection of studies and data extraction

After performing the literature search, we initially retrieved around 500 articles based on the keywords. From these, 350 articles were excluded after title/abstract review and 119 after full-text evaluation based on our inclusion/exclusion criteria. A total of 34 studies were identified for data extraction; however, only 33 studies were selected for data extraction (**Figure 1**), as [46] has reported contradictory outcomes in the results and discussion sections (in results showing a negative correlation but in the discussion claiming a positive correlation). In the end, our meta-analysis cohort included 4565 cases and 4907 healthy controls from the selected articles. A detailed description of the included studies is provided in **Suppl. Table 1**. Out of the 33 selected studies, 84.8% were conducted in India, 9.1% in Pakistan, and 6.1% in Bangladesh. No data was available on Sri Lankan, Bhutani, Maldivian, and Nepali populations. Based on the three commonly used genetic models; co-dominant, dominant, and recessive, 63.6% studies had reported a positive correlation and 3% a negative correlation with the DD genotype under the recessive genetic model **(Suppl. Table 1)**. However, the pooled data showed high levels of heterogeneity (I^2^ =80%) under both recessive and dominant models (**Figure 2 and 3**). All the relevant data mentioned in the M&M section was extracted manually from each study wherever available. The STREGA quality check indicated that 90.9% of the pooled studies were of high quality (scoring between 15-19) and the remaining studies were of medium quality, scoring between 12 and 14. (**Supplementary File 1**)

**Figure 2:**
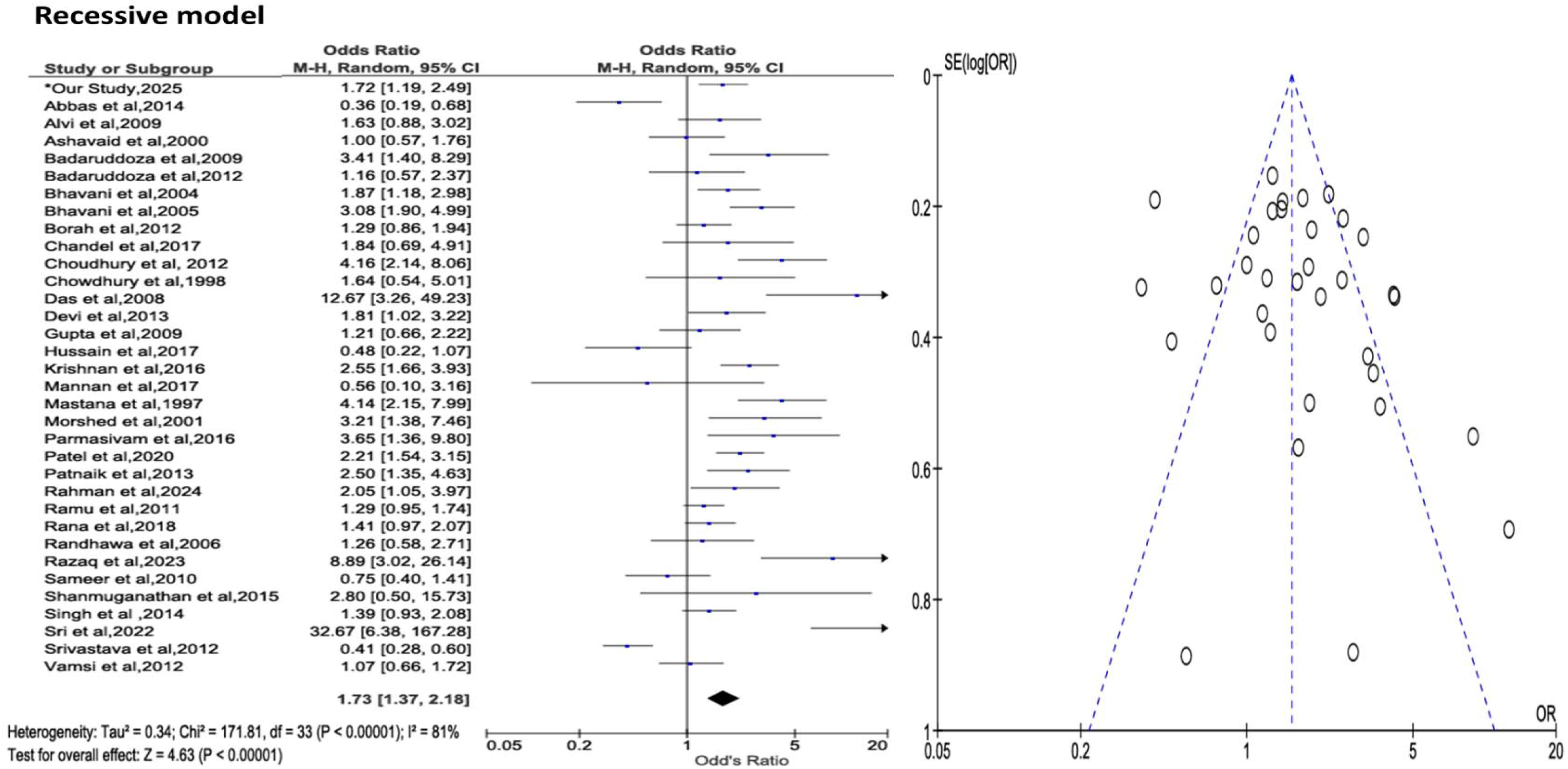
Forest plot of studies evaluating the relationship between ACE I/D polymorphism and essential hypertension under recessive model.

**Figure 3:**
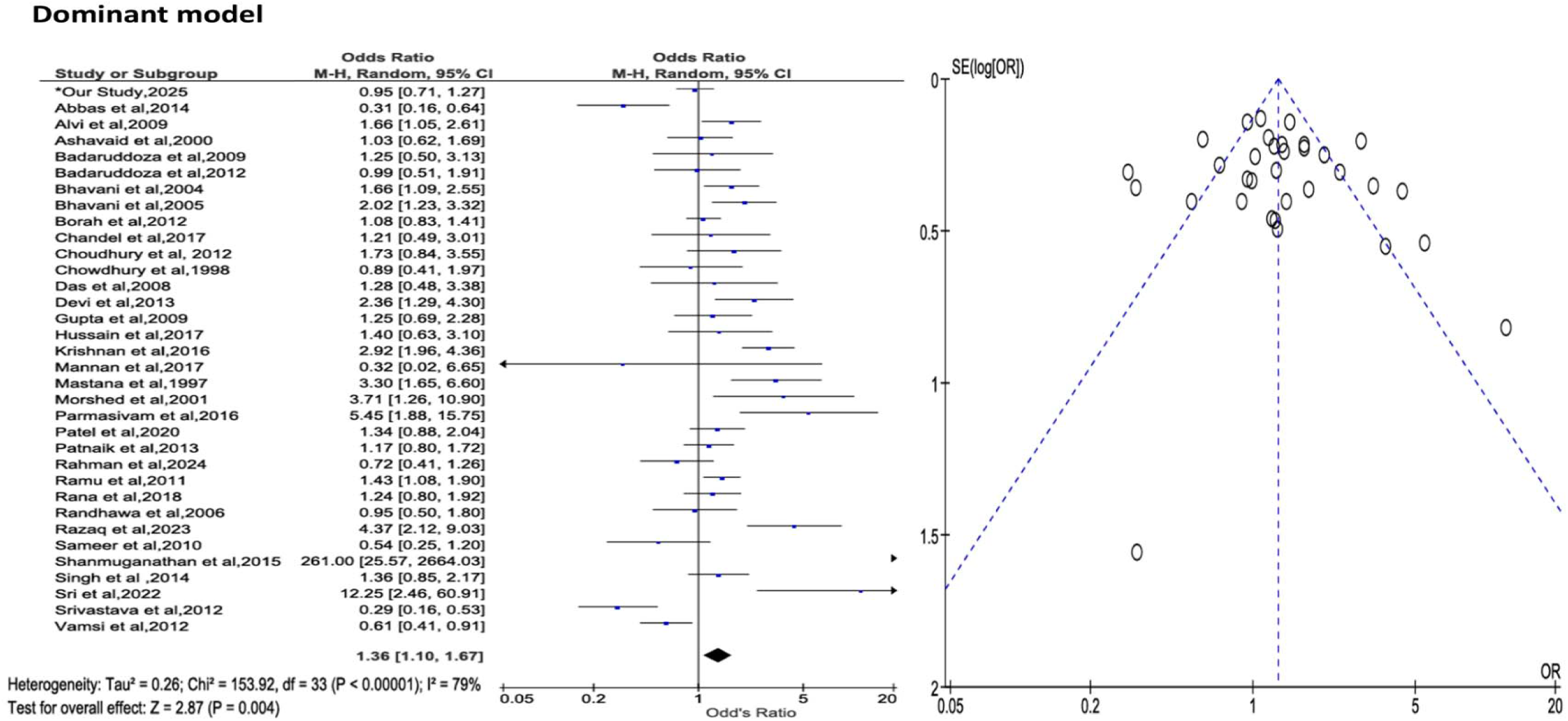
Forest plot of studies evaluating the relationship between ACE I/D polymorphism and essential hypertension under dominant model.

#### Statistical analysis

The statistical analysis was carried out on the pooled sample size of 10,289 subjects [4987 cases (4565 pooled + 422 own data) and 5302 controls (4907 pooled + 395 own data)]. The demographic and clinical characteristics of the pooled data are mentioned in the **Suppl. Table 6**. The percentage of males and females was 57.4% and 42.6% respectively. The average age of cases was 50.6±8.9 and controls 47.6±9.1. The allele and genotype frequencies of the cases and controls are given in **Table 4**. At the allelic level, no statistically significant differences were observed between cases and controls (P=0.31), similar to our case-control study. However, at the genotypic level, cases had a higher percentage of DD genotype than controls (29.5% vs. 22%) and a lower percentage of II genotype (28.6% vs. 34.1%). Calculated OR under the codominant and recessive genetic models revealed outcomes similar to our case-control study (**Table 4**). Under the recessive model, we observed a highly significant OR between the cases and controls [OR 1.48 (95%CI: 1.35–1.62), P<0.0001], suggesting a positive correlation of the DD genotype with EH. Likewise, under the codominant model, both II vs. ID and II vs DD comparisons revealed a positive correlation with EH [OR 1.13 (95%CI: 1.03–1.24, P=0.006), and OR 1.59 (95%CI: 1.43–1.76, P<0.0001), respectively]. Unlike our case-control study outcomes, a strong positive correlation was also observed under the dominant model (II vs. ID+DD) with an OR of 1.28 (95%CI: 1.18–1.40, P<0.0001) (**Table 4**).

**Table 4:**
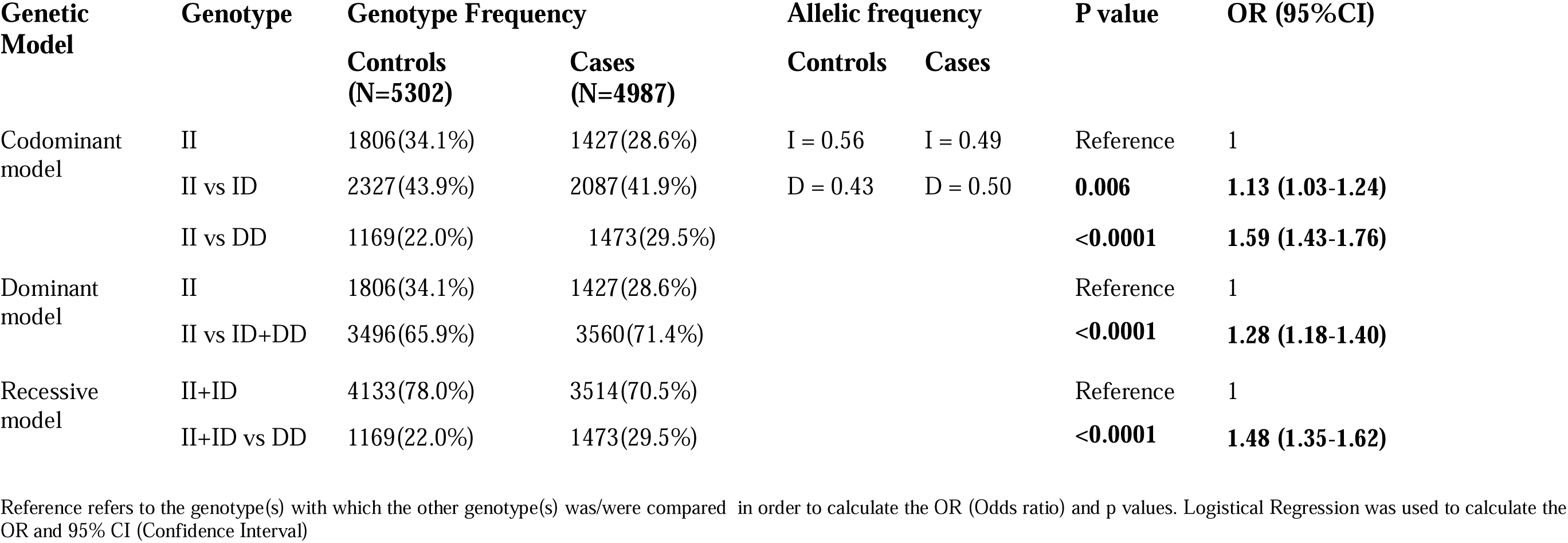
Genotype and allele frequency distribution in cases and controls under different genetic models and their statistical significance (Published + Our Study)

In the gender-wise analysis, we combined our data with the gender-wise genotype data from 11 available Indian studies[19, 24, 39, 42, 47-53]. Gender-wise data was not reported in of the other pooled studies. Statistical comparison was performed between 1313 male cases and 1425 male controls and between 923 female cases and 1013 female controls to investigate the relationship between EH and gender. In the whole data set, even though both males and females showed a significant positive correlation with *ACE* I/D, the OR and the non-adjusted P values were statistically strong in male (OR 1.79, P<0.0001) than in females (OR 1.38, P=0.003) under the recessive model (**Table 5**). Since both genders showed a positive correlation in the meta-analysis, we wondered if ethnicity has any role in the gender-specific risk that we observed in our North Indian population. To answer this, we split our gender data into South and North Indian groups and repeated the same statistical analysis. Interestingly enough, we observed a clear difference in gender susceptibility in the North Indian populations, with only men being at a 3-fold increased risk of EH [OR 3.10 (95%CI: 2.11-4.55), P<0.0001) if carrying the DD genotype (**Table 5**). These outcomes are in concordance with our case-control outcome, showing a clear distinction in susceptibility between the North Indian males and females. On the other hand, the South Indian population also showed gender wise differences in susceptibility with only males being at an increased risk of developing EH but only under the recessive model [OR 1.47 (95%CI: 1.18-1.84), P=<0.0001) (**Table 5**). However, the South Indian men were at lower odds of developing EH (OR 1.47) than their North Indian counterparts (OR 3.10). Other subcategory analyses, such as DBP, SBP, HTN Stage-I, and Stage-II were not performed due to the non-availability of such data in the published studies.

**Table 5:** Gender-wise association with essential hypertension in cases and controls under different genetic models (Pooled + Our Study)

| Genetic Model | Genotype | Males |  | P value | OR (95%CI) | Females |  | P value | OR(95%CI) |
| --- | --- | --- | --- | --- | --- | --- | --- | --- | --- |
|  |  | Controls | Cases |  |  | Controls | Cases |  |  |
| **Pooled + Our Study** |  | **N=1425** | **N=1313** |  |  | **N=1013** | **N=923** |  |  |
| Codominant model | II | 490(34.4%) | 394(30.0%) | Reference | 1 | 368(36.3%) | 276(29.9%) | Reference | 1 |
|  | II vs ID | 681(47.8%) | 551(42.0%) | 0.94 | 1.00 (0.84-1.19) | 444(43.8%) | 411(44.5%) | **0.04** | **1.23 (1.00-1.51)** |
|  | II vs DD | 254(17.8%) | 368(28.0%) | **<0.0001** | **1.80 (1.46-2.21)** | 201(19.8%) | 236(25.6%) | **<0.0001** | **1.56 (1.22-1.99)** |
| Dominant model | II | 490(34.4%) | 394(30.0%) | Reference | 1 | 368(36.3%) | 276(29.9%) | Reference | 1 |
|  | II vs ID+DD | 935(65.6%) | 919(70.0%) | **0.01** | **1.22 (1.04-1.43)** | 645(63.7%) | 647(71.0%) | **0.003** | **1.33 (1.10-1.61)** |
| Recessive model | II+ID | 1171(82.2%) | 945(72.0%) | Reference | 1 | 812(80.2%) | 687(74.4%) | Reference | 1 |
|  | II+ID vs DD | 254(17.8%) | 368(28.0%) | **<0.0001** | **1.79 (1.49-2.15)** | 201(19.8%) | 236(25.6%) | **0.003** | **1.38 (1.12-1.71)** |
| **South India** |  | **N=819** | **N=804** |  |  | **N=565** | **N=586** |  |  |
| Codominant model | II | 265(32.4%) | 218(27.1%) | Reference | 1 | 199(35.2%) | 170(29.0%) | Reference | 1 |
|  | II vs ID | 359(43.8%) | 332(41.3%) | 0.32 | 1.12 (0.89-1.41) | 237(41.9%) | 259(44.2%) | 0.07 | 1.27 (0.97-1.67) |
|  | II vs DD | 195(23.8%) | 254(31.6%) | **<0.0001** | **1.58 (1.22-2.05)** | 129(22.8%) | 157(26.8%) | **0.02** | **1.42 (1.04-1.94)** |
| Dominant model | II | 265(32.4%) | 218(27.1%) | Reference | 1 | 199(35.2%) | 170(29.0%) | Reference | 1 |
|  | II vs ID+DD | 554(67.6%) | 586(72.9%) | **0.02** | 1.28 (1.03-1.59) | 366(64.8%) | 416(71.0%) | **0.02** | **1.33 (1.03-1.70)** |
| Recessive | II+ID | 624(76.2%) | 550(68.4%) | Reference | 1 | 436(77.2%) | 429(73.2%) | Reference | 1 |
| model | II+ID vs DD | 195(23.8%) | 254(31.6%) | <0.0001 | **1.47 (1.18-1.84)** | 129(22.8%) | 157(26.8%) | 0.12 | 1.23 (0.94-1.61) |
| **North India + Our Study** |  | **N=473** | **N=353** |  |  | **N=374** | **N=268** |  |  |
| Codominant | II | 171(36.2%) | 104(29.5%) | Reference | 1 | 135(36.1%) | 91(33.9%) | Reference | 1 |
| model | II vs ID | 255(53.9%) | 159(45.0%) | 0.87 | 1.02 (0.74-1.40) | 171(45.7%) | 113(42.2%) | 0.91 | 0.98 (0.68-1.40) |
|  | II vs DD | 47(9.9%) | 90(25.5%) | <0.0001 | **3.14 (2.05-4.83)** | 68(18.2%) | 64(23.9%) | 0.13 | 1.39 (0.90-2.15) |
| Dominant | II | 171(36.2%) | 104(29.5%) | Reference | 1 | 135(36.1%) | 91(33.9%) | Reference | 1 |
| model | II vs ID+DD | 302(63.8%) | 249(70.5%) | **0.044** | **1.35 (1.00-1.82)** | 239(63.9%) | 177(66.0%) | 0.57 | 1.09 (0.79-1.52) |
| Recessive | II+ID | 426(90.1%) | 263(74.5%) | Reference | 1 | 306(81.8%) | 204(76.1%) | Reference | 1 |
| model | II+ID vs DD | 47(9.9%) | 90(25.5%) | <0.0001 | **3.10 (2.11-4.55)** | 68(18.2%) | 64(23.9%) | 0.07 | 1.41 (0.96-2.07) |

Reference refers to the genotype(s) with which the other genotype(s) was/were compared in order to calculate the OR (Odds ratio) and p values. Logistical Regression was used to calculate the OR and 95% CI (Confidence Interval)

## Discussion

Essential hypertension is a complex health condition with no specific cause(s) and a major risk factor for heart failure, kidney damage, and stroke [54]. Besides multiple well-known non-genetic risk factors, a strong genetic heritability has been linked with HTN in multiple family/twin and GWAS studies [9, 37, 55-59]. Numerous identified genetic factors include variations in genes involved in the salt metabolism pathways [9]. The most prevalent concept in the etiology of HTN that has emerged over several decades of research signifies the importance of interaction between multiple genetic and environmental factors along with ethnicity. The majority of the South Asian populations, including Indians are at a higher risk of developing HTN, with most countries having a prevalence of >20% [60]. The genetic predisposition in these populations remains largely unknown barring few studies on variants of some selected genes, importantly the RAS pathway genes[18, 24, 40]. The *ACE* I/D polymorphism has been identified as a risk factor for cardiovascular diseases, diabetes, infertility, and susceptibility to COVID-19, besides EH in many ethnic populations, including South Asian populations[26, 61-65]. The D allele linked high risk for EH and other diseases is presumed largely due to the high levels of serum ACE in DD individuals, thus the presence of a more active RAS[18, 24]. A higher risk of HTN with excessive salt intake and increased sensitivity to sodium intake, besides the increased vasoconstriction effect of elevated angiotensin-II are some of the outcomes of a more active RAS pathway [18, 24].

In this study, we are reporting outcomes from an original case-control study on the North Indian population of Jammu and a meta-analysis on the pooled data of 10,289 subjects (published+own data) of South Asian countries on the association of *ACE* I/D polymorphism with EH. The identification of ‘D’ as the risk allele in Jammu population (OR 1.68, 95% CI 1.16–2.43) is broadly in concordance with the South Asian data [34, 35, 47, 66, 67]. A significant positive correlation of DD genotype with both SBP (OR 1.68, 95%CI: 1.16–2.42) and DBP (OR 1.52, 95%CI: 1.04–2.20) independently, further highlights the potential role of D allele in EH in the study population. Our results from gender-based subgroup analysis were however the most surprising, showing a clear distinction in *ACE* I/D polymorphism associated risk in males and females. Males with DD genotype were at a ∼3-fold increased risk of developing EH than males with II genotype (P=0.001). Similar outcomes under the recessive genetic model further reiterate that the DD genotype is a risk factor for EH only in men (OR 2.68, 95%CI: 1.58–4.53). The non-significant differences in the genotype frequency between female cases and controls under all tested genetic models (**Table 3**), on the contrary, suggest a lack of association. Testing the possibility of any BP subgroup-associated correlation in both genders yielded outcomes similar to what was observed in the combined group (SBP+DBP), with males showing positive correlation with both SBP and DBP independently and females showing no association (**Table 3**). Likewise, in the stage-wise correlation analysis, I/D polymorphism showed strong (P=0.009) and mild (P=0.03) positive correlation with Stage-I EH under the recessive and co-dominant model but showed no correlation with Stage-II EH under any genetic model, except with ID genotype in the codominant model (P=0.01) (**Suppl. Table 5**). Another interesting finding suggests that instead of being a risk factor, the *ACE* I/D variant has a protective effect in females aged >45 years (OR 0.94, 95%CI: 0.52–1.70). However, this could be due to the small sample size, thus needs further validation. Among the subgroup outcomes, the gender-based outcomes in particular provided some interesting insights into the level of *ACE* I/D associated susceptibility in the studied population. Such divergence in susceptibility could be due to multiple reasons. First and foremost is the sexually dimorphic nature of BP due to biological gender differences and, importantly differences in the hormonal systems, with males usually at a higher risk of developing HTN than females [68, 69]. For instance, testosterone is known to activate the vasoconstriction arm of the RAS via the activation of ACE-Ang-II-AT1R pathway, whereas estrogen is known to induce vasodilation by upregulating the counterbalancing Ang-(1–7)-ACE2-MasR-AT2R pathway [70]. Thus, it is plausible that high levels of ACE in DD males will result in increased activation of ACE-Ang-II-AT1R pathway induced by testosterone, whereas the presence of estrogen will minimize the effect of D allele-mediated high ACE levels in females. The second potential cause could be differences in exposure levels to environmental risk factors, such as smoking and drinking, as females in South Asia rarely smoke and drink, mainly due to societal reasons. Smoking is a known activator of the sympathetic nervous system, which in turn affects the RAS [71]. Excessive alcohol consumption causes an imbalance in the RAS via oxidative stress [72], thus putting DD males at a higher risk of developing EH. The third possible reason is the small sample size, as we studied only 157 controls/175 cases in the female group and 238 controls/247 cases in the male group. These possibilities, along with any other confounding factor(s) will be studied in future research.

Our meta-analysis study outcomes were largely in line with our case-control study, supporting the D allele as the risk allele in the South Asian populations (**Table 4**). All three genetic models demonstrated a statistically highly significant correlation with EH in the pooled data set (**Table 4**). This is in agreement with the published data, as the majority of the studies we pooled data from have reported a moderate to strong association of *ACE* I/D with EH under different genetic models. The reason for the lack of association reported in some South Asian populations [35-37] could be primarily due to a smaller sample size, gender differences, ethnicity, or variations in exposure levels to environmental risk factors.

Outcomes similar to ours in the gender-wise meta-analysis, especially in the North Indians further reiterate the gender bias in the *ACE* I/D-associated EH susceptibility. Men with the DD genotype appear to be clearly at a significantly higher risk of EH than females. Whether a similar trend holds true for other South Asian populations; however, needs to be studied. Even though the biological role of this polymorphism can be presumed from the evidence showing high ACE expression in the presence of the D allele, direct evidence is still lacking. Additionally, more *ACE* polymorphisms may further impact its function, resulting in varied outcomes in different individuals. In future studies, we intend to measure the serum ACE levels in cases and controls to confirm whether the ACE levels indeed vary between the three genotypes and genotype more *ACE* polymorphisms in our study population.

## Conclusion

This study provides strong evidence in support of a positive correlation between the *ACE* insertion/deletion polymorphism and essential hypertension in the South Asian population, with the D allele being the risk allele. Ours is the first report providing convincing evidence in support of a differential susceptibility in males and females, with the DD genotype carrying men of Indian origin, particularly North Indians being at a higher risk of developing essential hypertension than females.

## Data Availability

All data produced in the present work are contained in the manuscript

## Acknowledgement

AB is the recipient of the ICMR-CAR grant (5/10/15/CAR-SMVDU/2018-RBMCH) and ICMR-EMR grant (6719/2020-DDl/BMS). The authors acknowledge Dr. Fayaz Ahmed Wani, Dr. Anupama Shah, and Dr. Pankaj Sharma from Govt. Medical College Jammu and Dr. Dharminder Kumar from Rama Krishna Medical Centre, Jammu, Jammu and Kashmir for their valuable contribution by facilitating sample collection and BP measurement. We are also thankful to the volunteers who participated in this study.

## Authors’ contributions

S.S. and S.B. generated and analyzed the data. S.S wrote the first draft. S.S and A.B. reviewed the pooled studies. S.B. reviewed the pooled studies when needed. A.D. revised and reviewed the manuscript. AB conceived the study, analyzed the data, revised and reviewed the manuscript.

## Ethics approval and consent to participate

Ethics approval and consent to participate: This study was approved by the Central University Jammu Human Ethics Committee (notification no. IHEC/CUJ/CMB-23/01). Written and informed consent was taken from all the participants.

## Availability of data and materials

All data generated and used and/or analyzed in this study is included in the manuscript.

---

**Suppl. Table 1:** Overview of basic characteristics information of included studies in meta-analysis.

| S. No | Population | Total no. of Individuals |  | Age |  | Gender |  |  |  | BMI |  | AVERAGE |  | AVERAGE |  | Reported Association | Reference |
| --- | --- | --- | --- | --- | --- | --- | --- | --- | --- | --- | --- | --- | --- | --- | --- | --- | --- |
|  |  | Cases | Controls | Cases | Controls | Males |  | Females |  | Cases | Controls | SBP |  | DBP |  |  |  |
| Pakistan |  |  |  |  |  |  |  |  |  |  |  |  |  |  |  |  |  |
| 1. | Khyber Pakhtunkhwa | 121 | 60 | 58 ± 7.2 | 55.4 ± 5.1<sup>#</sup> | 54 | 36 | 67 | 24 | 24.3 ± 3.1 | 28.1 ± 5.2 | 153.91 ±12.65 | 118.20 ±17.13 | 92.9± 5.7 | 74.12± 7.58 | Positive Association | (26) |
| 2. | Faisalabad | 100 | 48 | 52 ± 11 | 47 ± 8 | NA | NA | NA | NA | 26.3± 5.7 | 24.3 ± 4.3 | 147 ± 23 | 111 ± 15 | 90 ± 12 | 72 ±10 | Positive Association | (27) |
| 3. | Lahore (Punjabi) | 226 | 119 | 52.8 ± 9.8 | 57.4 ± 10.05<sup>#</sup> | 87 | 61 | 139 | 58 | NA | NA | 152.44 ± 12.3 | 116.24 ± 13.24 | 92.49 ± 6.72 | 73.55 ± 6.13 | No Association | (28) |
| Bangladesh |  |  |  |  |  |  |  |  |  |  |  |  |  |  |  |  |  |
| 4. | Dhaka | 44 | 59 | 47.3 ±11.5 | 43.5 ± 12.6 | 30 | 40 | 14 | 19 | NA | NA | 149.4 ± 6.21 | 118 ± 8.73 | 111.2 ± 9.53 | 82.1 ± 5.73 | Positive Association | (34) |
| 5. | Bangladesh | 51 | 52 | 46.3± 10.3 | 34.4± 10.8<sup>#</sup> | NA | NA | NA | NA | 25.5± 3.0 | 23.4± 3.6 | 139.0 ±17.9 | 110.9± 14.5 | 90.6± 9.1 | 76.7± 6.9 | No Association | (29) |
| India (North) |  |  |  |  |  |  |  |  |  |  |  |  |  |  |  |  |  |
| 6. | Uttar Pradesh | 138 | 116 | 41.2± 11.39 | 40.03 ±10.28 | 76 | 63 | 62 | 53 | 100.9 ±68.1 | 23.34± 3.0 | 147.6 ±21.16 | 131.8 ±128.3 | 88.85 ±11.0 | 78.66 ± 7.3 | Positive Association | (18) |
| 7. | New Delhi | 222 | 252 | 51.6± 7.2 | 49.7 ± 10.4 | 142 | 194 | 80 | 58 | NA | NA | 145.5 ± 14.05 | 120 ± 3.47 | 93.6± 8.0 | 80.49± 2.5 | I allele Positive Association | (30) |
| 8. | Kashmir | 52 | 150 | NA | NA | 38 | 100 | 14 | 50 | NA | NA | NA | NA | NA | NA | No | (37) |

|  |  |  |  |  |  |  |  |  |  |  |  |  |  |  |  | Association |  |
| --- | --- | --- | --- | --- | --- | --- | --- | --- | --- | --- | --- | --- | --- | --- | --- | --- | --- |
| 9. | Punjab | 100 | 100 | 52.8±<br>11.6 | 50.7±<br>10.07 | 40 | 40 | 60 | 60 | NA | NA | NA | NA | NA | NA | Positive<br>Association | (31) |
| 10. | Pathankot and<br>Ludhiana | 51 | 19 | 54.20<br>±1.5 | 51.05±<br>3.07 | NA | NA | NA | NA | NA | NA | 174.31<br>±5.03 | 122.00<br>±3.38 | 93.73<br>±1.45 | 86.74±<br>2.10 | No<br>Association | (20) |
| 11. | Haryana | 106 | 110 | 53.9<br>±13.9 | 51.96 ±<br>16.7 | 63 | 67 | 43 | 43 | 27.47<br>±3.42 | 23.38 ±<br>3.78 | 142.9<br>±5.87 | 117.69<br>± 3.62 | 92.44<br>±<br>4.60 | 75.63 ±<br>4.49 | No<br>Association | (2) |
| 12. | Himachal<br>Pradesh and<br>Punjab | 49 | 49 | 55.02<br>±13.5<br>2 | 49.87<br>±9.67 | NA | NA | NA | NA | 26.9±<br>4.27 | 25.93±<br>3.54 | 146.5±<br>17.64 | 130.12<br>± 13.22 | 90.32<br>±<br>10.06 | 79.97±<br>9.09 | No<br>Association | (22) |
| 13. | Punjab | 90 | 91 | 52.3±<br>6.54 | 52.3±<br>5.01 | 47 | 47 | 43 | 44 | 27.05<br>±3.08 | 26.36±<br>2.84 | 147.38<br>±11.49 | 131.46<br>±11.49 | 92.53<br>±10.3<br>5 | 79.50±<br>9.05 | No<br>Association | (33) |
| 14. | Punjab | 59 | 311 | NA | NA | 31 | 160 | 28 | 151 | NA | NA | NA | NA | NA | NA | Positive<br>Association | (19) |
| 15. | Punjab | 50 | 50 | 54.25<br>±<br>9.03 | 54.29<br>±18.42 | 28 | 28 | 22 | 22 | 29.32<br>±3.25 | 26.56 ±<br>13.19 | 148.23<br>± 9.11 | 129.31<br>± 10.13 | 92.14<br>±<br>7.37 | 85.19 ±<br>8.79 | Negative<br>Association | Badaruddo<br>za 2009 |
| 16. | Haryana | 206 | 245 | NA | NA | NA | NA | NA | NA | 18.35<br>±3.35 | 16.41<br>± 2.98 | NA | NA | NA | NA | Positive<br>Association | (35) |
| 17. | Gujarat | 279 | 292 | 49.2<br>±10.6 | 47.6 ±<br>9.2 | 181 | 169 | 98 | 123 | 25.4<br>± 3.8 | 24.6 ±<br>2.6 | 138.6 ±<br>10.4 | 129.8 ±<br>8.2 | 86.2<br>± 4.2 | 83.2 ±<br>2.8 | Positive<br>Association | (64) |
| **India (South)** |  |  |  |  |  |  |  |  |  |  |  |  |  |  |  |  |  |
| 18<br>. | Andhra Pradesh | 208 | 220 | 43.6±<br>5.6 | 42.78±<br>5.7 | 121 | 130 | 97 | 90 | 26.1±<br>3.2 | 25.3<br>±4.1 | 151<br>±9.8 | 123.34<br>±1.8 | 92.5±<br>8.7 | 69.89<br>± 5.0 | Positive<br>Association | (21) |
| 19<br>. | Andhra Pradesh | 100 | 100 | 58.9±<br>9.0 | 58.1±<br>9.0 | 80 | 77 | 20 | 23 | 25.7<br>± 3.2 | 24.6 ±<br>2.5 | 149.2 ±<br>9.1 | 130.3 ±<br>10.1 | 93.2<br>± 7.6 | 84.2 ±<br>8.5 | Positive<br>Association | Nirupama<br>devi<br>2013 |
| 20<br>. | Tamil Nadu | 211 | 211 | NA | NA | 109 | 130 | 102 | 81 | NA | NA | NA | NA | NA | NA | Positive<br>Association | (43) |
| 21<br>. | Tamil Nadu &<br>Pondicherry | 462 | 444 | 45.1<br>± 0.4 | 47.4 ±<br>0.4<sup>#</sup> | 228 | 201 | 234 | 243 | 23.0<br>± 0.3 | 22.9 ±<br>0.2 | 153.4 ±<br>0.8 | 117.5 ±<br>0.4 | 97.3<br>± 0.5 | 78.2 ±<br>0.3 | Positive<br>Association | (46) |
| 22<br>. | Tribal<br>populations | 33 | 37 | NA | NA | 13 | 17 | 20 | 20 | NA | NA | NA | NA | NA | NA | Positive<br>Association | (48) |
| 23<br>. | Hyderabad | 279 | 200 | 55.57<br>±9.78 | 47.63±<br>9.65<sup>#</sup> | 153 | 131 | 126 | 69 | 27.20<br>±4.73 | 25.80±<br>3.72 | 160.53<br>±20.98 | 120.05<br>±0.71 | 98.23<br>±12.8<br>7 | 80.0<br>±0.35 | II genotype<br>Positive<br>Association | (45) |
| 24<br>. | Hyderabad | 200 | 200 | NA | NA* | 146 | 147 | 54 | 53 | NA | NA* | NA | NA* | NA | NA* | Positive<br>Association | Bhavani et<br>al,2004 |
| 25<br>. | Chennai | 100 | 100 | NA | NA | NA | NA | NA | NA | NA | NA | NA | NA | NA | NA | Positive<br>Association | (32) |
| 26<br>. | Puducherry | 30 | 30 | 46.6<br>±5.40 | 39.83<br>±5.40 | NA | NA | NA | NA | NA | NA | NA | NA | NA | NA | Positive<br>Association | (22) |
| 27<br>. | Chennai | 101 | 81 | NA | NA | NA | NA | NA | NA | NA | NA | NA | NA | NA | NA | Positive<br>Association | (36) |
| 28<br>. | Tamil Nadu | 30 | 30 | 48.7±<br>12.14 | 42.14±<br>12.81 | 20 | 17 | 10 | 13 | NA | NA | 141±9 | 122±8 | 95±2 | 78±4 | Positive<br>Association | (23) |
| 29 | Mumbai | 105 | 192 | 50.2<br>±9.78 | 46.9<br>±10.77 | NA | NA | NA | NA | 27.21<br>±<br>4.56 | 23.50 ±<br>3.2 | 141.1<br>±14.43 | 122.01<br>± 10.1 | 89.0<br>±<br>7.88 | 81.56 ±<br>7.4 | No<br>Association | (14) |
| 30 | Hyderabad | 125 | 200 | 50.9±<br>0.94 | 45.06±<br>0.53 | NA | NA | NA | NA | 26.7±<br>0.43 | 25.4±0.<br>29 | 151.5±<br>2.28 | 152.8±<br>2.81 | 92.3±<br>0.90 | 93.1±<br>1.01 | Positive<br>Association | (24) |
| **India (East)** |  |  |  |  |  |  |  |  |  |  |  |  |  |  |  |  |  |
| 31 | Assam and<br>Mizoram | 407 | 509 | 44.5<br>±14.9 | 38.0 ±<br>14.5<sup>#</sup> | NA | NA | NA | NA | 21.1<br>± 4.2 | 21.1 ±<br>3.8 | NA | NA | NA | NA | No<br>Association | (56) |
| 32 | Calcutta | 185 | 165 | NA | NA* | 105 | 79 | 80 | 86 | NA | NA* | NA | NA* | NA | NA* | Positive<br>Association | (33) |
| 33 | Odisha | 246 | 274 | 49.4±<br>10.38 | 48.8±<br>11.04 | 159 | 158 | 87 | 116 | 24.2±<br>3.99 | 23.1±<br>1.9 | 148.4<br>±18.40 | 116.2<br>±5.40 | 93.18<br>±9.94 | 78.14<br>±4.24 | Positive<br>Association | (44) |
| **Total** |  | **4766** | **5116** |  |  | **1951** | **2092** | **1500** | **1499** |  |  |  |  |  |  |  |  |

\*Gender wise data available only <sup>#</sup> p value < 0.05,significant

**Suppl. Table 2:** Baseline Characteristics of Controls and Cases on the basis of DBP vs. SBP and Gender wise.

| Characteristics | Controls | Cases | P value |
| --- | --- | --- | --- |
| **DBP** | **N=559** | **N=258** |  |
| Age (Yrs) | 54.60±12.5 | 56.02±12.8 | 0.135 |
| BMI | 24.32±3.9 | 24.88±4.6 | 0.074 |
| Gender | Female (n=214)<br>Male (n=345) | Female (n=118)<br>Male (n=140) | 0.05 |
| DBP | 74.91 ±7.2 | 94.70 ±5.3 | ≤0.0001 |
| **SBP** | **N=418** | **N=399** |  |
| Age (Yrs) | 53.98±12.6 | 56.18±12.5 | 0.012 |
| BMI | 24.57±3.8 | 24.42±4.4 | 0.609 |
| Gender | Female (n=169)<br>Male (n=249) | Female (n=163)<br>Male (n=236) | 0.902 |
| SBP | 115.89±8.7 | 157.13±9.8 | ≤0.0001 |
| **Males** | **N=238** | **N=247** |  |
| Age(Yrs) | 54.41±12.4 | 56.32±11.9 | 0.085 |
| BMI | 24.89±3.9 | 24.32±4.4 | 0.13 |
| SBP | 115.01±8.4 | 155.44±12.9 | <0.0001 |
| DBP | 72.1±5.9 | 89.2±8.5 | <0.0001 |
| **Females** | **N=157** | **N=175** |  |
| Age (Yrs) | 53.76±12.9 | 55.29±13.4 | 0.29 |
| BMI | 24.05±3.5 | 24.62±4.4 | 0.21 |
| SBP | 116.2±8.6 | 154.9±12.1 | <0.0001 |
| DBP | 72.8±6.4 | 89.4±9.1 | <0.0001 |

BMI, Body Mass Index; DBP, Diastolic Blood Pressure, SBP, Systolic Blood Pressure; Yrs, Years. Unpaired t-test was performed to calculate the probability (p) values

**Suppl. Table 3:**
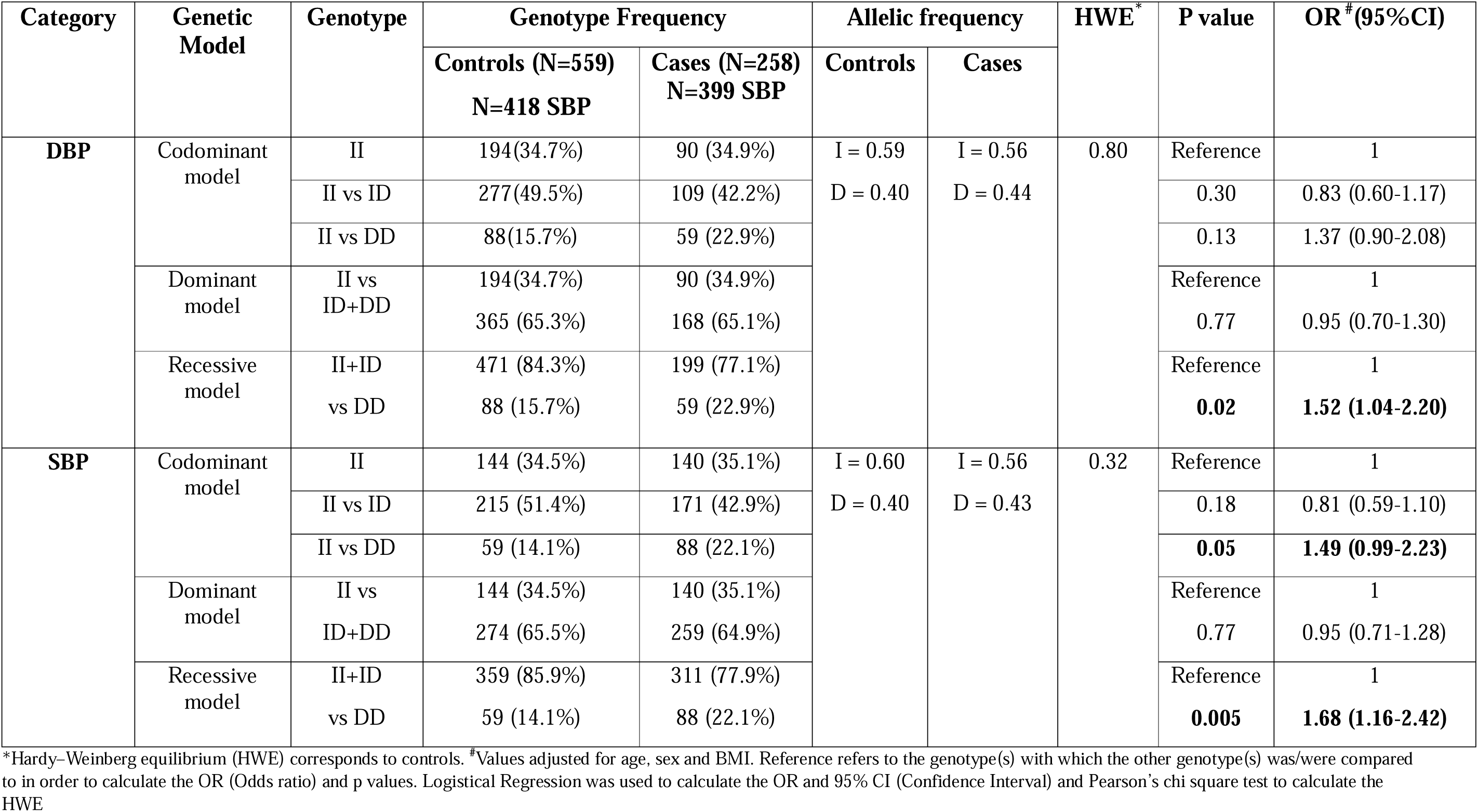
Genotype frequency and allelic frequency distribution under different genetic models on the basis of DBP vs SBP.

**Suppl. Table 4:** Genotype frequency distribution among cases and controls under different genetic models in the <45 and ≥45year age groups.

| Genetic Model | Genotype | Controls | Cases | P value | OR(95%CI) |
| --- | --- | --- | --- | --- | --- |
| **<45 Combined** |  | **N=104** | **N=65** |  |  |
| Codominant model | II | 39 (37.5%) | 23 (35.38%) | Reference | 1 |
|  | II vs ID | 56 (53.85%) | 32 (49.23%) | 0.21 | 1.95 (0.68-5.58) |
|  | II vs DD | 9 (8.65%) | 10 (15.39%) | 0.89 | 0.95 (0.48-1.88) |
| Dominant model | II vs | 39 (37.5%) | 23 (35.38%) | Reference | 1 |
|  | ID+DD | 65 (62.5%) | 42 (64.62%) | 0.80 | 1.08 (0.56-2.08) |
| Recessive model | II+ID | 95 (91.35%) | 55 (84.61%) | Reference | 1 |
|  | vs DD | 9 (8.65%) | 10 (15.39%) | 0.15 | 2.01 (0.76-5.30) |
| **Males** |  | **N=60** | **N=35** |  |  |
| Codominant model | II | 24 (40.0%) | 15 (42.9%) | Reference | 1 |
|  | II vs ID | 33 (55.0%) | 14 (40.0%) | 0.44 | 0.70 (0.28-1.74) |
|  | II vs DD | 3 (5.0%) | 6 (17.1%) | 0.14 | 3.19 (0.66-15.2) |
| Dominant model | II vs | 24 (40%) | 15 (42.9%) | Reference | 1 |
|  | ID+DD | 36 (60%) | 20 (57.1%) | 0.82 | 0.90 (0.38-2.14) |
| Recessive model | II+ID | 57 (95%) | 29 (82.9%) | Reference | 1 |
|  | vs DD | 3 (5%) | 6 (17.1%) | 0.07 | 3.85 (0.86-17.1) |

| Females |  | N=44 | N=30 |  |  |
| --- | --- | --- | --- | --- | --- |
| Codominant model | II | 15 (34.1%) | 8 (26.7%) | Reference | 1 |
|  | II vs ID | 23 (52.3%) | 18 (60%) | 0.46 | 1.55 (0.47-5.08) |
|  | II vs DD | 6 (13.6%) | 4 (13.3%) | 0.41 | 1.98 (0.38-10.1) |
| Dominant model | II vs | 15 (34.1%) | 8 (26.7%) | Reference | 1 |
|  | ID+DD | 29 (65.9%) | 22 (73.3%) | 0.39 | 1.63 (0.52-5.1) |
| Recessive model | II+ID | 38 (86.4%) | 26 (86.7%) | Reference | 1 |
|  | vs DD | 6 (13.6%) | 4 (13.3%) | 0.58 | 1.48 (0.35-6.17) |
| ≥45 Combined |  | N=291 | N=357 |  |  |
| Codominant model | II | 96 (32.99%) | 126 (35.30%) | Reference | 1 |
|  | II vs ID | 148 (50.86%) | 149 (41.73%) | 0.14 | 0.77 (0.54-1.09) |
|  | II vs DD | 47 (16.15%) | 82 (22.97%) | 0.18 | 1.35 (0.86-2.12) |
| Dominant model | II vs | 96 (32.99%) | 126 (35.30%) | Reference | 1 |
|  | ID+DD | 195 (67.01%) | 231 (64.70%) | 0.57 | 0.91 (0.65-1.26) |
| Recessive model | II+ID | 244 (83.85%) | 275 (77.03%) | Reference | 1 |
|  | vs DD | 47 (16.15%) | 82 (22.97%) | **0.02** | **1.57 (1.05-2.35)** |
| Males |  | N=178 | N=212 |  |  |
| Codominant model | II | 64 (35.9%) | 67 (31.6%) | Reference | 1 |
|  | II vs ID | 94 (52.9%) | 95 (44.8%) | 0.86 | 0.96 (0.61-1.50) |
|  | II vs DD | 20 (11.2%) | 50 (23.6%) | **0.006** | **2.38 (1.28-4.44)** |
| Dominant model | II vs ID+DD | 64 (35.9%)<br>114 (64.1%) | 67 (31.6%)<br>145 (68.4%) | Reference<br>0.37 | 1<br>1.21 (1.79-1.84) |
| Recessive model | II+ID vs DD | 158 (88.8%)<br>20 (11.2%) | 162 (76.4%)<br>50 (23.6%) | Reference **0.002** | 1<br>**2.44 (1.39-4.29)** |
| **Females** |  | **N=113** | **N=145** |  |  |
| Codominant model | II | 32 (28.3%) | 59 (40.7%) | Reference | 1 |
|  | II vs ID | 55 (48.7%) | 54 (37.3%) | **0.02** | **0.51 (0.29-0.91)** |
|  | II vs DD | 26 (23%) | 32 (22%) | 0.21 | 0.65 (0.33-1.28) |
| Dominant model | II vs ID+DD | 32 (28.3%)<br>81 (71.7%) | 59 (40.7%)<br>86 (59.3%) | Reference<br>**0.03** | 1<br>**0.56 (0.33-0.95)** |
| Recessive model | II+ID vs DD | 87 (77%)<br>26 (23%) | 113 (78%)<br>32 (22%) | Reference<br>0.85 | 1<br>0.94 (0.52-1.70) |

Reference refers to the genotype(s) with which the other genotype(s) was/were compared in order to calculate the OR (Odds ratio) and p values. Logistical Regression was used to calculate the OR and 95% CI (Confidence Interval)

**Suppl. Table 5:** Genotype frequency distribution under different genetic models on the basis of different stages of blood pressure.

| Genetic Model | Genotype | Controls | Cases | P value | OR<sup>#</sup>( 95%CI) |
| --- | --- | --- | --- | --- | --- |
| **Stage-I** |  | **N=395** | **N=258** |  |  |
| Codominant model | II | 135(34.2%) | 83(32.2%) | Reference | 1 |
|  | II vs ID | 205(51.9%) | 118(45.7%) | 0.73 | 0.94 (0.65-1.34) |
|  | II vs DD | 55(13.9%) | 57(22.1%) | **0.03** | **1.66 (1.05-2.65)** |
| Dominant model | II vs | 135(34.2%) | 83(32.2%) | Reference | 1 |
|  | ID+DD | 260(65.8%) | 175(67.8%) | 0.60 | 1.09 (0.78-1.52) |
| Recessive model | II+ID | 340(86.1%) | 201(77.9%) | Reference | 1 |
|  | vs DD | 55(13.9%) | 57(22.1%) | **0.009** | **1.73 (1.14-2.61)** |
| **Stage-I Males** |  | **N=238** | **N=156** |  |  |
| Codominant model | II | 88(36.9%) | 47 (30.1%) | Reference | 1 |
|  | II vs ID | 127(53.4%) | 72 (46.2%) | 0.76 | 1.07 (0.67-1.69) |
|  | II vs DD | 23(9.7%) | 37 (23.7%) | **0.001** | **2.93 (1.55-5.52)** |
| Dominant model | II vs | 88(36.9%) | 47 (30.1%) | Reference | 1 |
|  | ID+DD | 150(63.1%) | 109 (69.9%) | 0.16 | 1.36 (0.88-2.10) |
| Recessive model | II+ID | 215(90.3%) | 119 (76.3%) | Reference | 1 |
|  | vs DD | 23(9.7%) | 37 (23.7%) | **<0.0001** | **2.81 (1.59-4.96)** |
| **Stage-I Females** |  | **N=157** | **N=102** |  |  |
| Codominant model | II | 47(29.9%) | 36 (35.3%) | Reference | 1 |
|  | II vs ID | 78(49.7%) | 46 (45.1%) | 0.25 | 0.71 (0.40-1.27) |
|  | II vs DD | 32(20.4%) | 20 (19.6%) | 0.49 | 0.78 (0.38-1.59) |
| Dominant model | II vs | 47(29.9%) | 36 (35.3%) | Reference | 1 |
|  | ID+DD | 110(70.1%) | 66 (64.7%) | 0.25 | 0.73 (0.42-1.25) |
| Recessive model | II+ID | 125(79.6%) | 82 (80.4%) | Reference | 1 |
|  | vs DD | 32(20.4%) | 20 (19.6%) | 0.88 | 0.95 (0.50-1.78) |
| **Stage-II** |  | **N=395** | **N=164** |  |  |
| Codominant model | II | 135(34.2%) | 68 (41.5%) | Reference | 1 |
|  | II vs ID | 205(51.9%) | 63 (38.4%) | **0.01** | **0.61 (0.40-0.91)** |
|  | II vs DD | 55(13.9%) | 33 (20.1%) | 0.58 | 1.15 (0.68-1.95) |
| Dominant model | II vs | 135(34.2%) | 68 (41.5%) | Reference | 1 |
|  | ID+DD | 260(65.8%) | 96 (58.5%) | 0.09 | 0.72 (0.49-1.05) |
| Recessive model | II+ID | 340(86.1%) | 131 (79.9%) | Reference | 1 |
|  | vs DD | 55(13.9%) | 33 (20.1%) | 0.09 | 1.51 (0.93-2.44) |
| **Stage-II Males** |  | **N=238** | **N=92** |  |  |
| Codominant model | II | 88(36.9%) | 36 (39.1%) | Reference | 1 |
|  | II vs ID | 127(53.4%) | 38 (41.3%) | 0.23 | 0.72 (0.42-1.23) |
|  | II vs DD | 23(9.7%) | 18 (19.6%) | 0.08 | 1.91 (0.92-4.00) |
| Dominant model | II vs | 88(36.9%) | 36 (39.1%) | Reference | 1 |
|  | ID+DD | 150(63.1%) | 56 (60.9%) | 0.70 | 0.90 (0.55-1.49) |
| Recessive model | II+ID | 215(90.3%) | 74 (80.4%) | Reference **0.01** | 1 |
|  | vs DD | 23(9.7%) | 18 (19.6%) |  | **2.29 (1.16-4.50)** |
| Stage-II Females |  | N=157 | N=72 |  |  |
| Codominant model | II | 47(29.9%) | 32 (44.5%) | Reference | 1 |
|  | II vs ID | 78(49.7%) | 25 (34.7%) | **0.01** | **0.44 (0.23-0.85)** |
|  | II vs DD | 32(20.4%) | 15 (20.8%) | 0.24 | 0.63 (0.29-1.36) |
| Dominant model | II vs | 47(29.9%) | 32 (44.5%) | Reference | 1 |
|  | ID+DD | 110(70.1%) | 40 (55.5%) | **0.02** | **0.50 (0.28-0.90)** |
| Recessive model | II+ID | 125(79.6%) | 57 (79.2%) | Reference | 1 |
|  | vs DD | 32(20.4%) | 15 (20.8%) | 0.93 | 0.97 (0.48-1.94) |

#Values adjusted for age and BMI. Reference refers to the genotype(s) with which the other genotype(s) was/were compared in order to calculate the OR (Odds ratio) and p values. Logistical Regression was used to calculate the OR and 95% CI (Confidence Interval)

**Suppl. Table 6:**
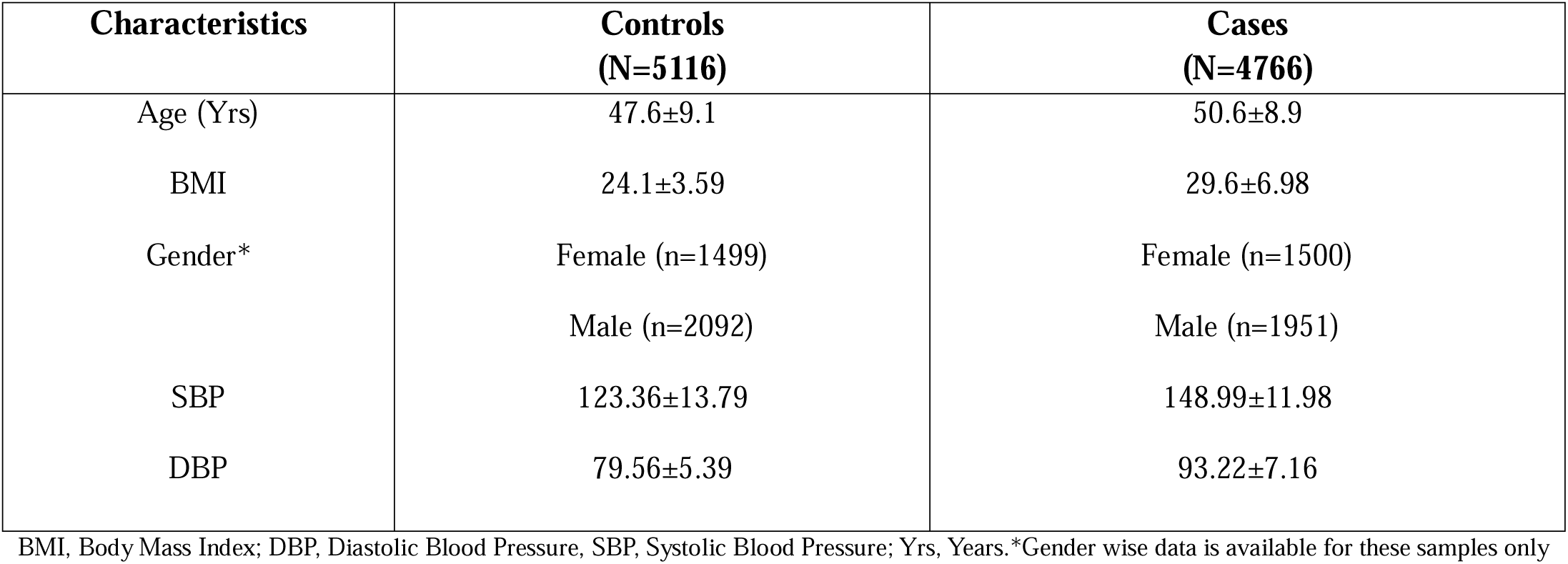
Demographic characteristics and clinical parameters of the controls and cases from pooled studies.

| **Characteristics** | **Controls**<br>**(N=5116)** | **Cases**<br>**(N=4766)** |
| --- | --- | --- |
| Age (Yrs) | 47.6±9.1 | 50.6±8.9 |
| BMI | 24.1±3.59 | 29.6±6.98 |
| Gender* | Female (n=1499) | Female (n=1500) |
|  | Male (n=2092) | Male (n=1951) |
| SBP | 123.36±13.79 | 148.99±11.98 |
| DBP | 79.56±5.39 | 93.22±7.16 |

BMI, Body Mass Index; DBP, Diastolic Blood Pressure, SBP, Systolic Blood Pressure; Yrs, Years.\*Gender wise data is available for these samples only

